# COVER-ME: Developing and Evaluating community-based interventions to promote vaccine uptake in East London minority ethnicity (ME) populations; underserved migrants and persons with low income: protocol for a pilot randomised controlled trial

**DOI:** 10.1101/2024.08.21.24312371

**Authors:** T. Chaudhry, P. Tum, Z H. Tam, A. Brentnall, H. Smethurst, K Kielmann, H. Kunst, S. Hargreaves, N J C. Campbell, C Griffiths, D Zenner

**Affiliations:** Wolfson Institute of Population, Queen Mary University of London, London, UK; Digital Environment Research Institute, Queen Mary University of London, London, UK; ApptHealth, Bolton, UK; Institute of Tropical Medicine, Antwerp, Belgium; Blizard Institute, Faculty of Medicine and Dentistry, London, UK; The Migrant Health Research Group, Institute for Infection and Immunity, City St George’s, University of London, London, UK; UK Health Security Agency, London, UK

**Keywords:** Vaccination, vaccine, vaccine uptake, Covid-19, Flu, underserved groups, migrant health, ethnic-minority, patient engagement

## Abstract

**Introduction:** Under vaccination amongst underserved groups remains low due to existing disparities. This is particularly the case with post-pandemic COVID-19 vaccinations, and other vaccine-preventable diseases including measles, Mumps and Rubella (MMR) or influenza. Therefore, we aim to 1) to determine the feasibility and practicality of implementing a patient engagement tool (PET) and gain vital insights to plan a subsequent definitive randomised controlled trial (RCT) to evaluate the effectiveness of this tool for increasing uptake of COVID-19 and Flu vaccination; 2) and to define the appropriate level of support needed for health care providers at site-level to ensure successful implementation of the PET and to identify supporting activities needed to implement interventions for COVID-19 and Flu vaccinations.

**Methods and Analysis:** This is a randomised controlled feasibility study evaluating a co-designed PET, involving randomisation at individual and cluster level. For individual randomisation, patients will be individually randomised 1:1 to receive the intervention (PET) or routine care; whereas for cluster randomisation six GP practices will be randomised 1:1, and divided into two tranches at two separate time points. Both groups will receive training and activation of the software. Data will be analysed using statistical software R (4.0 or greater) or STATA (17 or greater). Baseline characteristics will be summarised and presented in groups based on an intention to treat (ITT) basis with categorical data; including demographics, socioeconomic variables, co-morbidities, and vaccination status.

**Ethics and Dissemination:** Ethical approval was granted Westminster Ethics Committee (ref: 316860). Our dissemination strategy targets three audiences: (1) Policy makers, public and health service managers and clinicians responsible for delivering vaccines and infection prevention services; (2) patients and public from underserved population groups (3) academics.

## Background

Vaccinations can prevent up to 1.5 million deaths per year [1]; however, there are considerable disparities in uptake among some groups. These include migrants or socioeconomically deprived persons. Under-vaccination among these populations remains to be a problem in deprived regions of the UK, where there are barriers of implementing routine immunisation and vaccinations [2]. For example, during the COVID-19 vaccination roll out, vaccine uptake was considerably lower among Black and South Asian communities in comparison with the White British population, and was also lower among lower socioeconomic groups compared with the general population in the UK. Specifically, vaccine uptake has been low among black, Bangladeshi, and Pakistani communities who form a large proportion of the East London population [2]. Under-vaccination is also common for other vaccine-preventable diseases such as the measles, Mumps and Rubella (MMR) or influenza [1].

For such illnesses, including COVID-19, under-vaccinated population groups have higher risk of infection and disease and their under-vaccination can result in a higher burden of morbidity and mortality. For influenza, measles, and polio the picture is similar. This means that whilst the COVID-19 pandemic has ended and rates are decreasing, our research is transferrable and will guide current national vaccination efforts; to address under-vaccination overall and in specific population groups.

To explore vaccination uptake amongst marginalised groups, we conducted a qualitative study involving semi-structured interviews, focus group discussions and workshops with different community members and healthcare professionals within East London. This enabled us to collate social and cultural concepts, perceptions, and experiences and views for COVID-19 and other vaccines, across individuals of migrant and/or minoritised ethnic (ME) background. A systematic review was conducted to gain better insight of how different underserved populations vary in vaccination uptake behaviours; and to what extent and how certain determinants influence vaccination uptake in such groups. Factors identified that were associated with low-uptake of vaccine included mistrust in vaccine development, information (too much and/or too little), health and religious beliefs, travelling abroad, and frontline work situations, where working in the healthcare services or the public sector vaccine uptake was mandatory.

The qualitative study and systematic review informed the co-design of a feasible patient-engagement tool (PET) which is a digital platform available via a mobile phone; consisting of educational components such as informative text messages regarding the benefits of vaccine uptake, as well as reminders adapted to the needs and preferences of the target group to promote vaccination uptake.

For our detailed co-design we worked collaboratively with Appt Health [3], a service that can be used by healthcare commissioners and GP practices to engage patients to increase the uptake of preventive healthcare. We will then carry out a small pilot randomised controlled trial on the PET that has been developed for COVID-19 and Flu vaccination uptake for underserved at-risk populations including migrants and persons with ME backgrounds in East London.

With this pilot trial we aim to determine the feasibility and practicality of implementing the PET and gain vital insights to plan a subsequent definitive randomised controlled trial (RCT) to evaluate the effectiveness of this tool for increasing uptake of COVID-19 and Flu vaccination. We will also estimate COVID-19 and/or flu vaccination uptake and estimate the effect of the intervention, as well as measure vaccine uptake variation by population group, and other parameters which help us design the definitive RCT. In addition, we also aim to define the appropriate level of support needed for health care providers at site-level to ensure successful implementation of the PET and to identify supporting activities needed to implement interventions for COVID-19 and Flu vaccinations.

## Methods

### Overall Design

This is a randomised controlled feasibility study evaluating a co-designed PET. The PET was co-designed in the qualitative work packages with members of the community as well as healthcare professionals (Ethical approval: REF QMERC22.266) that precedes this feasibility trial. This feasibility study will involve individual and cluster level randomisation described below.

#### Individual randomisation

Randomisation will take place at appt-health level on behalf of each practice provide. Patients will be individually randomised 1:1 to receive the intervention (PET) or routine care. Appt-health is a nominated and appointed provider for the NHS and has data processing agreements in place with the East London GP practices. Appt-health therefore acts on behalf of patient-facing providers and this process has been used for numerous other health promotion activities.

Appt-health works as recognised provider with SLAs on behalf of the practices and this working relationship has taken place over many years for several (mostly public health related) programmes, including e.g., cervical screening. They have legitimate access to the data acting on behalf of the practices for public health programmes and hold robust data sharing agreements; this is approved at ICB level. Appt health will act on behalf of clinicians to identify the patients using our pre-defined criteria in our study also.

Individual randomisation will be stratified by GP, using a random block allocation list implemented into the software used for the study.

#### Cluster Randomisation

Six GP practices will be randomised 1:1, and divided into two tranches at two separate time points. This will include n=3 GPs being randomised first, and then later the other three practices. The start dates will be at least one month apart. This will enable additional comparison at cluster (practice level). The first group of GPs will receive training, activation of the software, and patients will be randomised to intervention or routine care workflows during the first study period. For the second group of GPs this will occur in the second period after randomisation, one month later.

During the first period, data will be used in all n=6 GP practices for a cluster comparison analysis. The individual-randomisation comparison will use data from all GPs, with the second group contributing data from the second period only. This individual and cluster randomised design will enable us to assess the feasibility of a study with both (or either) individual- and cluster-randomised comparisons

### Surveys

Patients’ views on the acceptability, user friendliness will be determined through a questionnaire which will be available through a link which will be sent out via a text message. The questionnaire will also be available in paper format and in local languages.

A healthcare provider questionnaire will also be distributed to evaluate the feasibility and acceptability of the PET among healthcare providers. These will be distributed by the research team in paper format completed by staff at each GP surgery.

#### Selection Criteria and Recruitment

Inclusion criteria are listed in box 1. We will operationalise eligibility of study participation based on age, gender, ethnicity, and postcode and whether they are able to receive text messages, based on coding in the practice patient information system (Egton Medical Information Systems (EMIS) which provides patients full name and address).

Only individuals who consented to receive routine care messages from their GP surgery are enrolled and they will receive a message with an option to decline study participation.

#### Inclusion criteria

- Patient registered at study site (GP practice)
- Adult (aged 18y or older) at time of randomisation
- Eligible for COVID-19 vaccination and/or Flu vaccine (ie. Not received either a first, second or booster vaccination)

AND

From an underserved population group, defined

(1) non-white ethnicity

OR

(2) resident in a postcode in the bottom **20%** of index of multiple deprivation

OR

(3) Those receiving low income based on postcode of residence

#### Exclusion criteria

- They are unable or unwilling to consent (including those who do not consent to text messaging; those who opt out from taking part in research studies).

#### Gaining Patient Consent

Eligible participants will have consented to receive routine care messages from their GP surgery and have an option to decline participation. EMIS codes will be used to determine eligibility of study participation based on age, gender, ethnicity, and postcode.

Participants will receive a message containing opt-out consent. The process of consent has been split up into three parts below:

1. Consent to receive messages from the PET-Those with an EMIS code have consented to receiving messages from the GP and PET
2. Consent to use data (anonymously) – Participants will be sent a text message to opt-out if they refuse to have data shared.
3. Consent for Survey- a message for opt-in consent will be sent. The first part of the survey covers consent and data sharing procedures to which participants will agree or disagree to.

Regarding data sharing, all data will be completely anonymised for analysis (including stripping PID, such as NHS number, address, names, DOB etc).

Appt Health have a Data Processing Agreement in place with every practice they work with which covers the legal basis for contacting patients. This data sharing is a routine entity which has been utilised for multiple health promotion and preventative care projects and is fully compliant with GDPR and GCP.

#### Outcome measures

##### Primary outcome

The primary outcome is vaccination uptake in patients individually randomised. This will be measured as the number (percent) of relevant SNOMED codes in eligible patients. Uptake will be measured from the time that the eligible group in each practice is identified (and randomised) until 6 months follow-up (>180 days since randomisation).

##### Secondary outcome

1. Vaccination uptake in patients after 3- and 9-months follow-up (>90 and >270 days). This will be measured as the number (percent) of relevant SNOMED codes in eligible patients identified during the period.
2. Mean vaccination rate after 3-, 6- and 9-months follow-up. This will be measured as the number of patients vaccinated divided by the follow-up time. Time is defined as time from randomisation until the earliest of vaccination, leaving the GP, withdrawal the consent of use of data, or death.
3. Acceptability of the intervention To patients:
  a. Proportion of patients randomised to the intervention who engage with the PET and / or linked patient resources, as determined by user statistics logged on the software for
    i. The SMS messaging tool (number of SMS sent).
    ii. Number (percent) of patients who view the linked patient awareness resources.
    iii. Usage of the patient work list tool, which consist of specific targets for example, vaccine status and patient lists from the specified GPs included in the study.
  b. Patients’ responses of the questionnaire about the acceptability and user friendliness. To staff:
    a. Number and proportion of eligible patients randomised to the intervention processed using the patient work list tool.
    b. Healthcare providers’ responses of the questionnaire to evaluate the feasibility and acceptability of the PET among healthcare workers.
    c. The feasibility of study-related work processes, using process observations.
4. Feasibility of the intervention and randomisation
  a. Number and proportion of eligible patients randomised
  b. Support needed for healthcare providers at site-level for implementation (using process observations to understand and analyse environment and workflows).
  c. Clinical capacity: number of slots available for vaccination bookings for each GP practice and each day, by appointment time
5. Feasibility of the study design for a subsequent trial
  a. Number and proportion of patients with all inclusion / exclusion data available on the electronic health records
  b. Number and proportion of patients eligible for the intervention
  c. Number and proportion of patients randomised who are sent information via text messages from the intervention (letter / sms)
  d. Number and proportion of patients eligible by vaccination status (none, first, second, with booster or without booster)
  e. Number and proportion of patients who opt out for their data to be used or withdraw from the study
  f. Number and proportion of patients who consent for further questionnaires
  g. Number and proportion of patients who are booked for a vaccination appointment
  h. Number and proportion of patients who are categorised a “failed encounter” (not booked for appointment, no more action taken)

#### Sample size

In a given GP practice size of 10,000 patients, we estimate at least 40% meet the eligibility criteria based on their ethnicity or deprivation quintile. If 90% of these are already vaccinated, then at least n=400 patients will be eligible for randomisation at each GP, therefore, a total of 2400 eligible patients from six GP practices will be randomised 1:1. If 10% of these patients would get vaccinated over 6 months without an intervention, then our study would have approximately 90% power to show a 40% increase in vaccination rate (from 10% to 14.4%) when testing between the two groups using a z-test at the 5% level. Whilst we expect loss to follow up to be small, we will report the number of participants who withdraw from the study after randomisation.

### Analysis

All analysis will be undertaken using the statistical software R (4.0 or greater) or STATA (17 or greater). We will summarise baseline characteristics including demographics, socioeconomic variables, co-morbidities and vaccination status.

Characteristics will be presented in groups based on an intention to treat (ITT) basis with categorical data as number and percentage and continuous data as mean/ standard deviations or median/ inter-quartile range.

In the primary analysis, we will present uptake of COVID-19 and Flu vaccination in each arm on intention to treat (ITT) basis. We will assess the individual-level-randomised component. This analysis will include all patients who are eligible for the intervention, and who were randomised to receive one of two workflows on an individual basis (standard of care or the PET). All patients from first group of GPs (n=3) in both study periods, and only patients from second group (n=3) who are eligible in the second period (i.e., any eligible patient from first period in the second group of GPs who was vaccinated in the first period would not be part of the analysis sample) will be included. Uptake (from time of randomisation) will be estimated overall. Uptake 95% CIs will be obtained based on binomial assumption, as well as from a mixed-effects model that allows for hierarchical (random effects) variation by GP and booster groups. Logistic regression with adjustment described below will be used to estimate the marginal odds ratio of the PET compared with standard of care, and a profile-likelihood ratio confidence interval for the odds ratio will be reported. Adjustment of prior COVID-19 and influenza vaccination status, GP practice, age and sex will be used for the logistic regression model.

Secondary endpoints will be reported as point estimates with 95% CIs as appropriate. Summary statistics for each outcome by arm will be presented on an intention-to-treat basis. Categorical data will be shown with number and percentage. Continuous data will be shown as median and inter-quartile range, or mean, standard deviation if approximately normal.

Analysis of the primary and secondary endpoints and potential heterogeneity in uptake by prespecified subgroups will be undertaken. This includes GP practice, deprivation, age group, sex, and ethnicity.

We will explore the cluster-randomised component of this study. This analysis of uptake will take clustering in the design into account by using generalised linear models with normal random intercepts, and the model will also be used to obtain an estimate of the intra-class correlation coefficient. CIs will be based on methods that are most suitable when the number of clusters is not large. We will also explore whether a per-protocol analysis would be possible, by determining feasibility to define compliance / contamination from individual randomised allocation using process data.

#### Missing Data

Most of the data will be complete by design, and data capture maximised where feasible. If the level of missing data is likely to affect reported point estimates and estimates of effect size then multiple imputation will be used for analyses when appropriate, as well as complete-case analysis.

#### Assessment and Management of Risk

Although, this research is unlikely to inflict any risk or harm, the participant may not want to share or disclose their vaccination status. It is stressed to only share information a participant feels comfortable sharing. However, if participants feel distressed or do not wish to respond to certain questions asked during surveys, they may take a break and return when they are ready. The research team can signpost them to their local healthcare provider or to easily accessible services, such as mental health charity services across various sites in UK providing counselling for psychological support. Participants would be directed to access additional support from their care provider for referral to other agencies if issues raised.

All participants are eligible to withdraw at any point during the study if they wish to, without affecting on-going healthcare. They will also be given the opportunity to have their data fully withdrawn.

#### Annual Safety Reporting

The chief investigator (CI) (DZ) will send an Annual Progress Report to the REC and the sponsor using the Health Research Authority (HRA) template on the anniversary of the REC “favourable opinion”.

#### Data management

Electronic case report forms will be used. Routine data and vaccination status will be extracted from the patient’s electronic patient record. This database will be stored on a secure folder using a bespoke database from CASTOR, access will be limited to data administrators and investigators. A full audit trail will be generated of amendments to the database.

#### Record Retention and Archiving

All records are the responsibility of the CI and must be kept in secure conditions. When the trial is complete, it is a requirement of the Research Governance Framework and Trust Policy that the records are kept for a further 25 years. For trials involving Barts Health Trust patients, undertaken by Trust staff, or sponsored by Barts Health Trust or QMUL, the approved repository for long-term storage of local records is the Trust Modern Records Centre (Barts Health NHS Trust, 9 Prescot Street, London, E1 8PR 9 Prescot Street).

#### Monitoring and Auditing

On site monitoring will be performed as per the study monitoring plan. Monitoring will include source data verification to ensure data integrity as well as compliance to the protocol and GCP. The sponsor or delegate retains the right to audit any study, study site, or central facility. Any part of the study may be audited by the funders, where applicable.

#### Trial Committees

The day-to-day management of the study will be carried out by a Study Management Group (SMG), supported by a Patient and Public Involvement (PPI) advisory group and a data monitoring committee. The SMG with overall responsibility of the study will be chaired by the main applicant (Dr Zenner) and membership will include co-applicants, key collaborators and a PPI representative. The SMG will meet fortnightly, holds responsibility for the overall management and ensures that all project deliverables and stages are completed to time and budget. We will identify a PPI advisory group through collaborations .The SMG will also liaise regularly with key external stakeholders, including Public Health England (vaccination and inclusion departments), local CCGs, GP networks and local authorities as well as with groups, who research related topics to exchange information on national and regional vaccination efforts.

#### Public Involvement

Our team is well connected to local health service and community organizations with excellent links to NHS England and Public Health England. For the proposed research we will closely follow NIHR (previously INVOLVE) guidance to ensure the research is done ‘with’ rather than ‘to’ local communities. To coordinate this, we have already reached out to local community organizations and will establish a Patient and Public Involvement (PPI) advisory group, who directly reports to the study SMG. We will make use of our existing networks to establish the PPI advisory group, which will help coordinating PPI involvement at all stages of the study.

We will work closely with patients, and local community groups at all stages of the project, particularly with representatives of key vulnerable groups in East London. The PPI advisory group will advise us on the development of the study protocol, the approach to participants, developing the ethics application and supporting material such as information sheets and posters. It will also be advising us on the conduct of interviews and focus groups which will include guidance on content of questions for the interview guide. Individuals from our PPI group will be asked to “walk through” the study before recruitment starts, to highlight potential questions that participants may have. PPI involvement will be central to any input into further development of the study and will be involved at every step. We are also planning to support training members of local underserved communities as community researchers in collaboration with Social Action for Health (SAfH), to particularly support the qualitative component that has already taken place (Ethical approval: REF QMERC22.266). We are also keen to develop local research capacity in the community, which we hope will stay as a legacy of this project.

#### Dissemination of Research Findings

A meeting will be held after the end of the trial to allow discussion of the main results among the collaborators prior to publication. The success of the trial depends entirely on the wholehearted collaboration of a large number of patients, carers and health care professionals. For this reason, chief credit for the main results will be given not to the committees or central organisers but to all those who have collaborated in the trial. A writing committee will be convened to produce publications on behalf of the Trial Steering committee and the Study Management Group, they will not be permitted to publish data obtained from participants in the trial that use study outcome measures without discussion with the Chief Investigator and/or the Trial Steering Committee. Only anonymized data will be used for dissemination of research findings.

Our dissemination strategy targets three audiences: (1) Policy makers, public and health service managers, and clinicians responsible for delivering vaccines and infection prevention services; (2) patients and public from underserved population groups (3) academics.

#### Trial registration

The trial has been registered with ClinicalTrials.gov (Trial ID: NCT05866237)

#### Ethics

Ethical approval was granted from Westminster - Research Ethics Committee (ref: 316860).

## Discussion

This protocol outlines the processes in the co-design, development, and piloting of an effective, acceptable, and feasible PET for COVID-19 and flu vaccination uptake for underserved at-risk populations in East London.

Vaccination uptake is a complex and multifaceted process. Our preceding qualitative research showed that reasons for poor uptake range from concerns about the vaccine itself (e.g. adverse effects) to practical and logistical health access barrier issues to issues around the availability and presentation of information. Multiple interacting factors influencing vaccine uptake have also been observed in a recent systematic review of vaccination uptake among migrants [2]. Findings have demonstrated that people from marginalised or minoritized groups maybe more inclined to trust familiar and accessible sources, including their GP, rather than more distant parts of the health system, like the Department of Health and Social Care (DHSC). Therefore, there is a clear need to ensure greater engagement of local primary care providers and suitable interventions to promote vaccination uptake.

Based on previous research [3,4] and lessons from our own qualitative data, we co-designed a patient engagement intervention to address low vaccine uptake in underserved population groups. This pilot RCT is designed to assess the feasibility and effectiveness of a PET in an underserved population group. It is a preparatory study for a larger RCT and explores multiple issues around feasibility, acceptability, and statistical uncertainty. We expect that our initial findings will inform a larger RCT to explore under-vaccination at much larger scale amongst other populations across the UK.

Our study has some limitations including the small sample size which may also be affected by opt-in consent, as patients declining to take part could reduce sample size, which could affect the power calculation. A smaller sample may also not allow us to explore certain variables, or conduct sub-group analyses. In addition, the cluster design may have limitations exploring the individual effects and feasibility of the PET. However, we will explore this further during individual and cluster randomisation, and we will analyse individual data in individual randomisation.

Our study is the first step to testing a co-designed patient engagement tool, which may mitigate low vaccine coverage, amongst underserved and minoritised communities in East London and similar groups across the UK.

## Data Availability

Data is not yet available as this is a study protocol for a future study

## Authors’ contributions

TC conceptualized the study aims, study design, inclusion, and exclusion criteria, and data curation. Senior support was provided by DZ, who is also the CI of this study. PT will be supporting the study by co-ordinating tasks and processes involved in recruitment of GP practices and patients for randomisation.

HZT and AB are the statisticians who will be assisting in the analyses of results. HS is founder of appt-health, and has assisted in the co-design of the PET. HS is also responsible for randomisation of patients.

TC wrote all drafts of the manuscript with input from DZ, all co-authors listed verified the analysis, commented on the manuscript, and approved for submission.

## Sponsor Contact Information

Joint Research Management office, Dept W, Research services 69-89 Mile End Road, E1 4UJ, Telephone 02078827207

